# The Brief Observation of Symptoms of Autism (BOSA): Development of a New Adapted Assessment Measure for Remote Telehealth Administration through COVID-19 and Beyond

**DOI:** 10.1101/2021.11.01.21265761

**Authors:** Deanna Dow, Alison Holbrook, Christina Toolan, Nicole McDonald, Kyle Sterrett, Nicole Rosen, So Hyun Kim, Catherine Lord

**Affiliations:** University of California, Los Angeles (UCLA) Semel Institute for Neuroscience and Human Behavior; 760 Westwood Plaza, Los Angeles, CA 90024; Simons Foundation Autism Research Initiative; 160 5th Ave, New York, NY 10010; Department of Psychiatry, Weill Cornell Medical College, New York-Presbyterian Hospital, 21 Bloomingdale Road, Rogers Building, White Plains, NY 10605, USA

**Keywords:** Autism spectrum disorder, diagnosis, assessment, screening

## Abstract

Interest in telehealth assessment for autism has increased due to COVID-19 and subsequent expansion of remote psychological services, though options that are easy for clinicians to adopt and available through the lifespan are limited. The Brief Observation of Symptoms of Autism (BOSA) provides a social context with standardized materials and activities that can be coded by clinicians trained in the Autism Diagnostic Observation Schedule (ADOS). The current project examined psychometric properties to determine optimal use for each BOSA version. Three hundred and seven participants with 453 BOSAs were included to determine best performing items for algorithms, validity, sensitivity, specificity, recommended cut-offs, and proposed ranges of concern. While preliminary, the BOSA provides a promising new option for telehealth-administered assessment for autism.

---

The COVID-19 pandemic sparked heightened interest in flexible screening and assessment measures for autism spectrum disorder (ASD) that can be conducted remotely via telehealth without requiring face-to-face administration by a clinician. While some attention toward telehealth or technology-based evaluation had begun prior to the pandemic (Dahiya et al., 2020; Duda et al., 2016; Tariq et al., 2018), research on the efficacy of these measures was still in its early stages. Additionally, logistical barriers, such as lack of insurance coverage in the United States for clinical telemedicine services, made offering these services in medical settings largely impractical. However, when clinical services and research protocols were halted due to the pandemic, the field was forced to quickly seek and adopt innovative approaches that were feasible through telehealth or in-person visits with necessary safety protocols, leading to rapid expansion of remote services and approval of insurance-based telehealth coverage. Even as pandemic safety protocols have started to loosen, there is continued interest in the use of flexible, remote approaches to ASD assessment, which likely will continue to shape how services are delivered going forward. Not only would expansion of services improve feasibility and convenience for families, but it could also improve access to care by allowing those in more remote areas to be seen without traveling out of their local area. Bolstering empirical evidence for newly developed tools and methods of assessment is imperative as the field moves to embrace these new approaches.

## Telehealth ASD Assessment Measures

Several measures that were developed pre-pandemic have been offered as potential remote assessment options. Some of the most widely used (Jang et al., 2021) include the TELE-ASD-PEDS (Corona et al., 2020), Systematic Observation of Red Flags of ASD (SORF; Dow et al., 2020), Naturalistic Observation Diagnostic Assessment (NODA; Smith et al., 2017), and the Childhood Autism Rating Scale (CARS-II; Schopler et al., 2010). While the TELE-ASD-PEDS and SORF offer promising options for young children under the age of three, the TELE-ASD-PEDS had only previously been studied in a laboratory setting and psychometric properties are not yet available for use in the home (Wagner et al., 2021), and research on the SORF has only been conducted on the full one-hour home observation by videographers with no data yet substantiating use of a shorter, more feasible timeframe. The NODA is also a promising option for children, though research assessing its specificity in differentiating ASD from non-ASD developmental delays has not yet been conducted. The CARS-2 offers an option that can be used through adulthood but has not yet been validated for use without live, in-person observation.

Despite having some options for remote assessment, many groups expressed interest in continuing to administer the Autism Diagnostic Observation Schedule, Second Edition (ADOS-2; Lord et al., 2012a, 2012b) with modifications, likely due to its widespread use and number of clinicians trained to administer it. The ADOS-2 also boasts a strong empirical base (Lebersfeld et al., 2021), the ability to assess individuals across the age span, and had previously been required by many insurance providers to ensure coverage of medical services. However, standard administration of the ADOS-2 was not possible during the pandemic, and the adaptations needed altered the social environment when administered in-person while wearing face masks or using barrier shields. Anecdotally, concerns about the impact of masks, both on the client and provider, have been supported by reports from clinicians and parents. Significant adaptations are required for telehealth administration as well, such as only asking social and emotional questions and engaging in conversations for verbally fluent individuals, or having caregivers administer prompts for less verbal individuals and/or young children. While there has been some support for the utility of Module 4 remote administrations in terms of sensitivity with an all ASD sample (Schutte et al., 2015), the specificity of the instrument using standardized scoring would likely be compromised given that the full span of activities cannot be administered to elicit certain behaviors of interest, and nonverbal communication is more difficult (or arguably impossible, in the case of eye contact) to read when directed to a screen. For younger children, requiring caregivers to prompt their children through ADOS-2 tasks changes the standardized way the activities are carried out. While a parent-mediated context may provide rich qualitative information, the standardized scoring from the ADOS-2 should not be applied. Without the ADOS-2 during the pandemic, however, there was a gap where the gold-standard tool to assess autism used to lie, which left many clinicians and researchers searching for a convenient and effective replacement when other available options did not seem like a good fit for their clinic population or set-up.

## Introduction to the Brief Observation of Symptoms of Autism (BOSA)

One benefit of the ADOS-2 is that it provides a relatively natural and consistent context for clinicians to observe social communicative behaviors and to place these observations within a framework of standardized codes. This makes it an important counterpart to caregiver-report or self-report of symptoms and behavior, which can provide an incomplete picture of an individual’s needs. With this in mind, the Brief Observation of Symptoms of Autism (BOSA; Lord et al., 2020) was developed to provide a similarly naturalistic social context with standardized materials and activities, adapted from the Brief Observation of Social Communication Change (BOSCC; <u>Grzadzinski</u> et al., 2016) and ADOS-2 (Lord et al., 2012a, 2012b). The BOSA consists of a 12-14 minute interaction between an individual and a caregiver or clinician. New materials and/or conversational contexts are presented every 2-4 minutes while the dyad interacts naturally together using the materials and/or prompts.

The administration of the four versions of the BOSA is based primarily on tasks employed in previously developed BOSCC versions (i.e., MV or “minimally verbal”, PSYF or “phrase speech-young fluent”, F1 and F2 for “fluent” 6 through 10 years olds (F1) and “fluent” 11-year-olds through adults (F2)), chosen according to the individual’s age, language, and developmental level. Materials were modified and selected to create a standardized social context consisting of ADOS-2 toys and materials, interactive games that elicit shared affect, and question cards for older children and adults that include conversation prompts and ADOS-2 questions related to emotional experiences, social relationships, and responsibility. There are two sets of toys or materials used in each administration – one for the first approximately 6 minutes and one for the second 6 minutes (with slight differences in timing for the different versions).

The BOSA-MV is for individuals of any age who are nonverbal, have single words or only rote phrases, and consists of two sets of ADOS-2 toys and bubbles. The BOSA-PSYF is intended for individuals of any age who use flexible phrase speech or for individuals who are verbally fluent under ages 6 to 8 years old and includes two sets of ADOS-2 toys, bubbles or a rocket launcher, and a dollhouse or toy mailbox to help elicit conversation about the materials. Toy sets include action figures, dolls, furniture, purse with accessories, Poppin’ Pals, ball, plates, a pinball game and other materials taken from the ADOS-2 kit. Between the ages of 6 and 8, clinical judgment should be used based on the child’s verbal ability and attention span, as the PSYF is more structured and play-based, versus the F1 which is more conversational.

The BOSA-F1 is for verbally fluent children as young as 6 and up through age 10 and involves turn-taking games, answering socioemotional and conversation-starter questions, and having two unstructured conversations without materials present. The BOSA-F2 is for children from age 11 through adults and involves similar activities as the F1 with more advanced, age-appropriate games as well as questions and conversations. A short game of Slap Jack or tabletop basketball is used as a “warm-up,” then games are played while asking and answering questions using question cards (many taken from the socio-emotional questions in the ADOS-2) that correspond to the color of the game piece. Recommended games include Pop the Pig for younger fluent individuals and Jenga for older fluent individuals, as well as other interactive games.

After administration, a clinician trained in the ADOS-2 who has observed the BOSA live or through videorecording, scores the appropriate ADOS-2 protocol based on the participant’s age and language level, as well as any additional codes provided in the BOSA manual for that module. Clinicians should not score items if they do not have enough information to make an accurate judgment regarding the presence or absence of a particular symptom. ADOS-2 scores are then transferred onto a DSM-5 Checklist broken down into ASD symptom categories and converted to binary BOSA scores to indicate presence or absence for each symptom. Additional columns are also provided to note evidence collected from parent report or other observations to aid in determining whether the individual meets diagnostic criteria across domains.

## Purpose of this Study

The current project aimed to examine the psychometric properties and optimal use of the BOSA, as supported by empirical evidence and data-driven scoring procedures. Converted ADOS-2 scores were examined by module for best-performing BOSA algorithm items, including sensitivity and specificity. Because of COVID-19 related restrictions, we were aware from the start that our immediate focus would be on sensitivity of the BOSA, because our ability to recruit comparison groups into a new study was very limited; we provide some data on specificity whenever we were able to obtain it. Cutoff scores for each algorithm and suggested ranges of concern were developed to aid in clinical utility, and convergent validity with the ADOS-2 was examined for modules with large enough samples. As with various versions of the ADOS and ADI, we consider this work preliminary and hope it will provide a basis for replications and likely revisions with larger and more representative samples.

## Methods

### Preliminary Analysis and Development of Coding System

Prior to creating the BOSA coding system, distributions of ADOS-2 scores were examined in a large existing database of well-documented individuals with ASD and those with related, but non-ASD disorders (ASD n=3027, non-ASD n=1177) to determine how to maximize sensitivity when scores are collapsed into a binary coding system. A binary system was chosen because the primary purpose of the BOSA is not to determine severity but simply to indicate whether the presence of an ASD symptom was observed, without ruling out that it might occur in other circumstances. Standard ADOS-2 items from each module were included, as well as selected items from other modules if there was perceived clinical value. Items that were highly correlated or difficult to code in the BOSA context (i.e., determined by at least 50% with a code of “8”, meaning not codable) were removed from analyses. Checklists based on Diagnostic and Statistical Manual, Fifth Edition (DSM-5; APA, 2013) criteria were created for each module, onto which ADOS-2 codes can be transferred. “Recode rules” to convert raw ADOS item scores to a binary coding system were created based on these results and provided on the DSM-5 Checklists for each module. For the BOSA, a score of 0 represents absence of a clinically significant symptom and a score of 1 represents presence of a clinically significant symptom. The checklists can be used to view symptom presence across each diagnostic domain. There are additional columns provided to add information gained from outside observation and caregiver/teacher report to complement the results from the BOSA and to assist with clinical impressions.

### Participants

This sample included 307 unique participants with 453 observations for participants ranging from age 15 months to 42 years (see Table 1 for Participant Demographics). The Toddler Module included 94 observations from 47 children (29 ASD, 21 females) with ages ranging from 15 to 38 months (M = 25.30, SD = 4.31). Module 1 included 59 observations from 37 children (34 ASD, 5 female) ranging from 31 to 84 months (46.19 months, SD = 14.12). Module 2 included 76 observations from 55 children (32 ASD, 9 female) ranging from 29 months to 11 years (M = 51.53 months, SD = 21.71 months). Module 3 included 163 observations from 117 participants (76 ASD, 33 female) ranging from 4.0 to 16.17 years (M = 6.53 years, SD = 2.45 years). Module 4 included 61 observations from 51 participants (35 ASD, 10 female) ranging from 14.08 to 42.08 years (M = 26.50 years, SD = 6.02 years). Participants were recruited from 4 sources: 1) individuals participating in research studies or clinical treatment programs at UCLA (n=207) including the Baby Brain Imagining and Behavior (Baby BIBS) study, Expressive Movement Initiative (EMI), JASPER Early Intervention for Tuberous Sclerosis Complex, the Early Diagnosis/Longitudinal study, KidsConnect partial hospitalization program, Children’s Friendship Program social skills program, and PEERS for Careers and PEERS for Dating social skills programs; 2) children participating in research at the Center for Autism and the Developing Brain (CADB) at Weill Cornell (n=208) including the Simons Verbal BOSCC study, Kindergarten School Readiness study, CADB Early Intervention study, and the Video Feedback Intervention study; 3) individuals seen for a comprehensive diagnostic assessment at the UCLA Child and Adult Neurodevelopmental (CAN) Clinic (n=20); and 4) non-ASD volunteers recruited as study control subjects (n=18). All participants received either a BOSA or BOSCC administration; the BOSCC largely corresponds to the BOSA in length of time, activities, and materials available, with a few minor changes in materials based on availability. All originally administered BOSCCs will be referred to as BOSAs going forward given the similarity between standardized administrations and utilization of the same coding definitions and protocol.

**Table 1.**
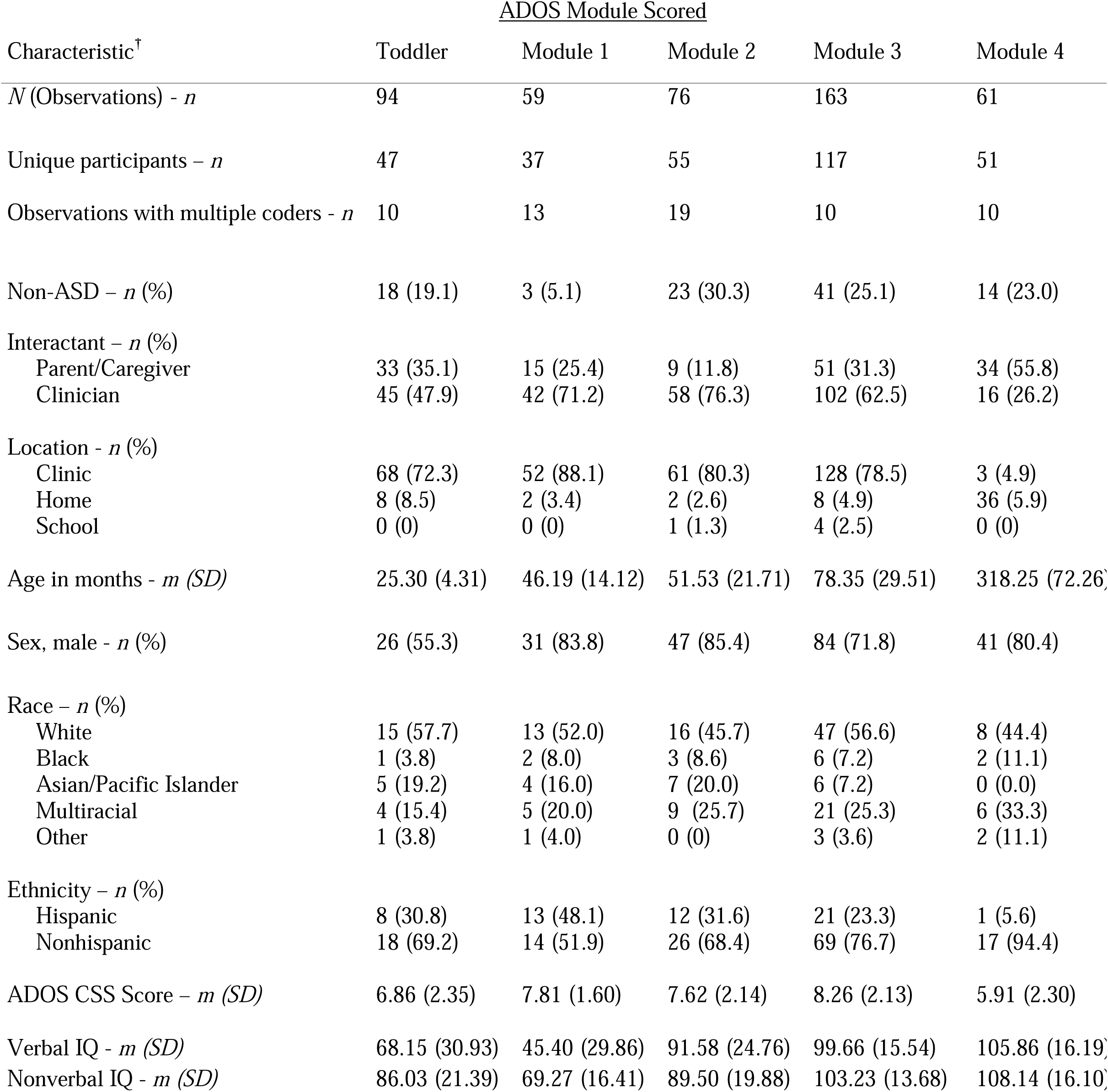
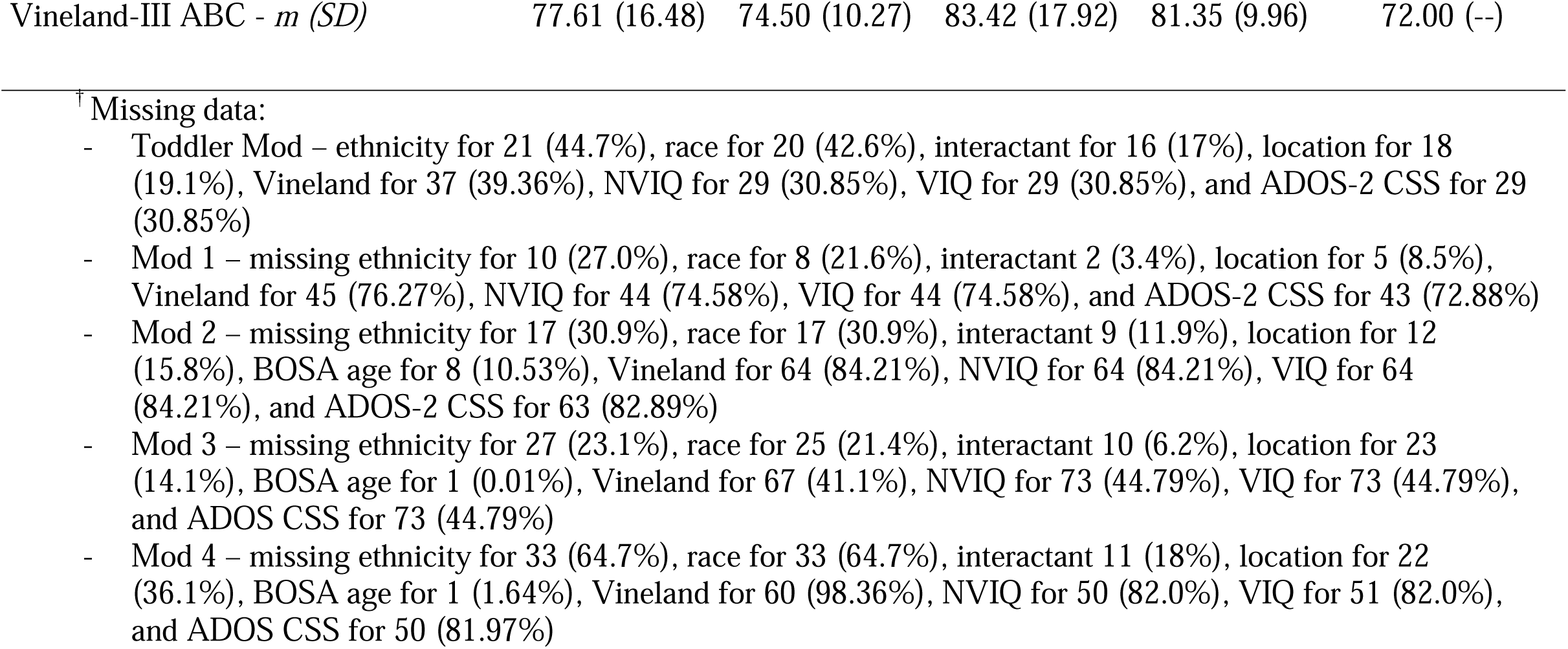
Participant Demographics

### Diagnostic Procedures

For all CADB participants, participants without a diagnosis participated in a comprehensive diagnostic assessment including an ADOS-2 and Autism Diagnostic Interview-Revised (ADI-R; Rutter et al., 2003). If they had been previously diagnosed with ASD by a medical provider, diagnosis was confirmed with an ADOS-2 but a full comprehensive diagnostic assessment was not required. Diagnoses given at UCLA’s autism clinic were determined based on all relevant information obtained during the evaluation, including the ADI-R, questionnaire data, collateral information, and behavioral observations from testing. Participants enrolled in treatment groups at UCLA often had a prior medical diagnosis of autism. A “best estimate” clinical or research diagnosis was determined for participants from UCLA research studies by a clinical psychologist or graduate level research associate with supervision by a licensed clinical psychologist with expertise in ASD-specific diagnostic assessment. “Best estimate” diagnoses used all relevant data, which, depending on the study or clinic protocol, included a developmental interview or the ADI-R and direct observation (using the BOSA administration and in some cases, an ADOS-2 administration). Thus, clinical diagnosis was not always independent of the BOSA; however, BOSA algorithms were derived after samples were collected and cutoff recommendations were not available at the time of determination. Participants with typical development (17.0%) and previously-established non-spectrum disorders (4.9%; e.g., developmental delay, anxiety disorders, attention-deficit/hyperactivity disorder) were included in the sample, though the majority was autistic (78.1%).

### Coding and Reliability Procedures

Both caregiver- and clinician-administered BOSAs were included in the analyses. Administrations done by caregivers were either done in-person (n=76) or through telehealth (n=20), with a research associate or clinician giving prompts for when to switch activities. Clinician BOSAs were completed prior to the pandemic. Percent agreement between caregiver and clinician BOSAs for a subsample of participants who completed both (n=46) was 86% within 3 points, 70% within 2 points, 47% within 1 point, and 23% exact agreement. Video recordings of the BOSAs were scored by graduate- or PhD-level research or clinical staff who were research reliable on ADOS-2 scoring and had established BOSA reliability. Coders established reliability by scoring three consecutive videos per module with at least 80% agreement with the master coder on item-level BOSA binary scores prior to beginning independent coding.

### Data Analytic Plan

A correlation matrix was constructed using the ADOS-2 items scored based on the BOSA observation. Items were removed if they correlated over .70 with any other item or if they were unable to be scored (receiving a score of 8 or N/A code) at least 50% of the time within each scoring module. The remaining items were used in the factor analysis to determine consistency of model fit with the two-factor model of the ADOS-2 and DSM-5 criteria for Social Affect (SA) and Restricted, Repetitive Behavior (RRB) domains.

Confirmatory Factor Analysis (CFA) was conducted for the BOSA items within each ADOS coding module using all available data to optimize sample size, including scores from children at multiple timepoints and videos coded by multiple coders. Analyses were conducted using Mplus (Muthén & Muthén, 1998-2011). Participant was included as a cluster-level unit to take into account multiple codings across participants. Items with significant loadings onto the SC and RRB factors were included in the algorithms, with the exception of low frequency items that were observed in fewer than 50% in ASD participants in this context. Cutoffs were determined in order to prioritize sensitivity around 90% while maintaining adequate specificity (though specificity data were limited due to the small non-ASD sample).

Receiver operator characteristic (ROC) curves were run for each module on the algorithm total, Social Affect total, and Restricted, Repetitive Behaviors total to determine how well the measure differentiates between ASD and nonspectrum groups. Sensitivity provides the proportion of individuals who are correctly identified as having ASD, while specificity shows the proportion of individuals who are correctly identified as nonspectrum. Area under the curve (AUC) shows the strength of discrimination between groups. ROC curve results were only reported for algorithm totals, as domain-specific algorithms are considered to be preliminary.

Ranges of concern were also identified to improve clinical utility, given our concerns about the preliminary nature of these data. Consistent with the ADOS-2 Toddler Module (Luyster et al., 2009) and the ADI-R algorithm for toddlers and preschoolers (Kim et al., 2013), three ranges are provided: Little-to-no concern, mild-to-moderate concern, and moderate-to-severe concern. Ranges of concern were determined by examining distributions of algorithm scores in ASD versus nonspectrum groups and were set so that 90-95% of participants with ASD would fall into one of the two groups suggesting clinical concern, with consideration to reduce the number of nonspectrum participants that would fall into the concern range.

Most participants in the current sample did not have an ADOS administration, with less than 10 completed each for Modules 1, 2, and 4. Despite the limited data, we felt it was important to include convergent validity results for modules that had enough participants with both BOSA and ADOS-2 administrations to obtain interpretable results (i.e., the Toddler Module and Module 3). Given that the BOSA has been used in place of the ADOS-2 during the pandemic, understanding the convergence between the measures for those modules gives some preliminary perspective on the clinical interpretation of scores. Pearson correlation coefficients were calculated between the BOSA algorithm total score, ADOS-2 algorithm total score and calibrated severity score (CSS), given the continuous nature of total scores.

Interrater reliability was assessed using a subsample of 30 videos, 10 each from MV, PSYF, and F1/F2 combined across available coders, with 10% of videos (i.e., 3 out of 30 videos) across sites (i.e., UCLA and CADB). For reliability analyses, items that could not be scored were removed and the remaining items were assessed using intraclass correlation coefficients (ICCs).

The administrations were randomly selected and coded by coders blind to diagnosis. Test-retest reliability was completed on 10 participants who completed two BOSAs within a 1-week time period. Five of the 10 participants had a diagnosis of ASD.

## Results

### Confirmatory Factor Analysis

CFA results supported good fit across modules using a 2-factor model of Social Affect and Restricted, Repetitive Behaviors, consistent with ADOS-2 algorithm subdomains and the DSM-5 (see Table 2). The Comparative Fit Index (CFI) ranged from .94 to .97, Tucker-Lewis Index (TLI) ranged from .93 to .96, and Root Mean Square Error of Approximation (RMSEA) ranged from .05-.14. Item factor loadings (see Table 3) ranged from .40 to .93 for Toddler Module, .25 to .98 for Module 1, .50 to .95 on Module 2, .55 to .96 on Module 3, and .40 to .99 for Module 4. The lowest factor loadings were RRBs, consistent with what has been reported for the ADOS-2 (Gotham et al., 2007, 2008) and BOSCC (Grzadzinski et al., 2016).

**Table 2.** Confirmatory Factor Analysis Fit Results

|  | CFI | TLI | RMSEA |
| --- | --- | --- | --- |
| Toddler | .96 | .96 | .05 |
| Module 1 | .94 | .93 | .09 |
| Module 2 | .95 | .94 | .07 |
| Module 3 | .97 | .96 | .06 |
| Module 4 | .96 | .95 | .14 |

CFI=Comparative Fit Index; TLI=Tucker-Lewis Index, RMSEA=Root Mean Square Error of Approximation

**Table 3.**
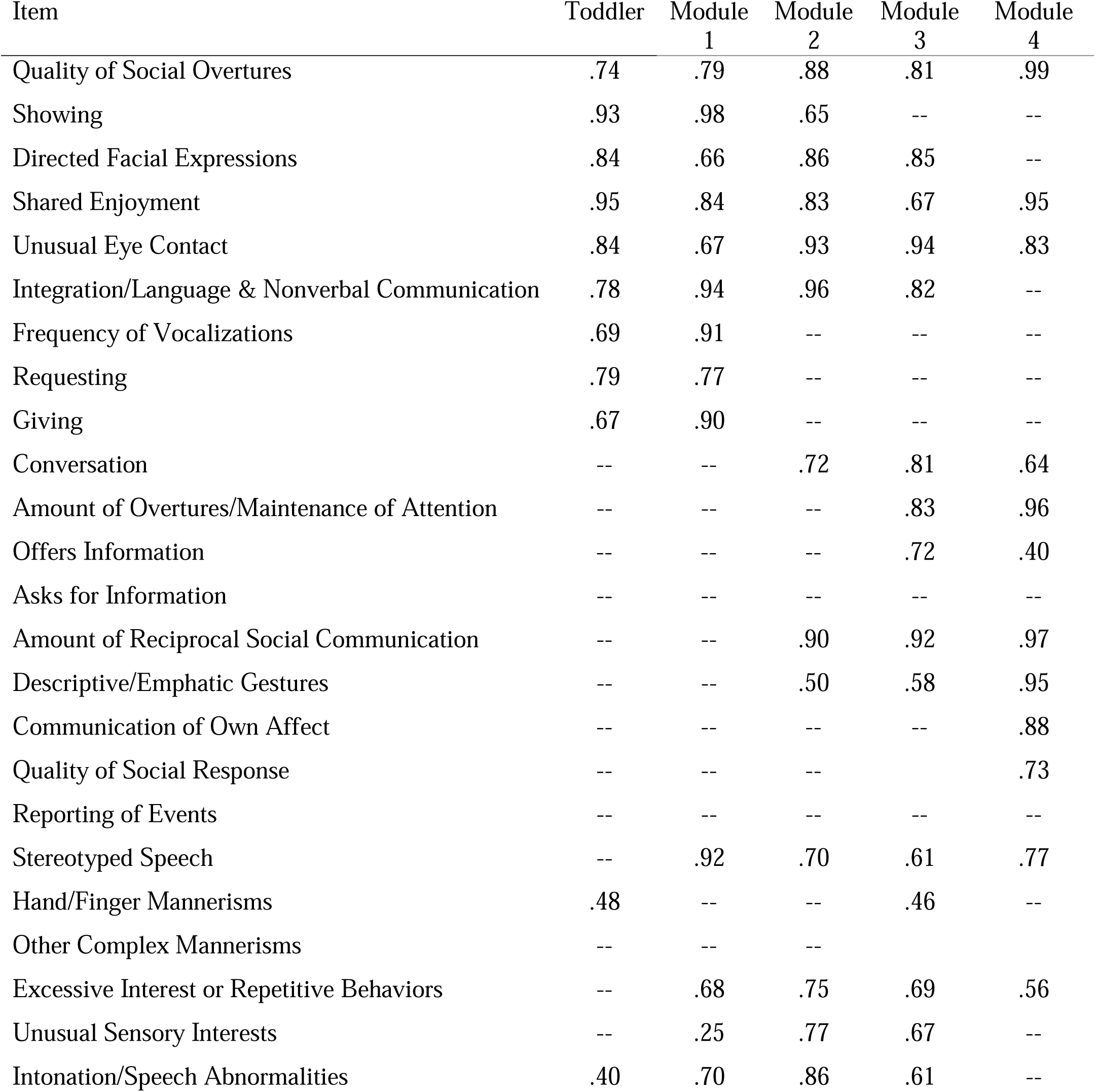
Algorithm Item Factor Loadings by Module

### ROC Curve Testing and Ranges of Concern

Total algorithms resulted in good discrimination between ASD and nonspectrum groups (see Tables 4 and 5; Figure 6 for ROC curve results). For the Toddler Module scoring, AUC was .96, sensitivity was 96% and specificity was 83% at a recommended cutoff of 6. Ninety-six percent of the ASD group scored in the mild-to-moderate or moderate-to-severe concern groups. The Module 1 sample resulted in similar discrimination (AUC=.97), sensitivity of 91% and 100% specificity (though the non-ASD sample was very small) at a recommended cutoff of 5. Due to the small non-ASD sample and therefore limited data to support specificity, a higher cutoff was used to keep sensitivity right around 90%. Ninety-three percent of participants with ASD fell in one of the two concern groups. Module 2 scoring resulted in an AUC of .87, sensitivity of 91%, specificity of 74%, and a cutoff of 9; 98% of ASD participants fell into one of the concern groups. AUC for Module 3 was .91, sensitivity was 86%, and specificity was 70% with a cutoff of 6 and 91% of ASD participants within a concern group. Module 4 resulted in high discrimination (AUC=.98), sensitivity (91%), and specificity (100%) with a cutoff of 3, with 92% of the ASD group falling into a concern range.

**Table 4.** Total Algorithm ROC Curve Results, Recommended Cutoffs and Ranges of Concern

|  | AUC | Sensitivity | Specificity | Recommended Cutoff | Range of concern |
| --- | --- | --- | --- | --- | --- |
| Toddler | .96 | 96% | 83% | 6 | 0-3 Little-to-No<br>4-5 Mild-to-Moderate<br>6+ Moderate-to-Severe |
| Module 1 | .97 | 91% | 100% | 5 | 0-4 Little-to-No<br>5-8 Mild-to-Moderate<br>9+ Moderate to Severe |
| Module 2 | .87 | 91% | 74% | 9 | 0-6 Little-to-No<br>7-8 Mild-to-Moderate<br>9+ Moderate-to-Severe |
| Module 3 | .91 | 86% | 70% | 6 | 0-3 Little-to-No<br>4-5 Mild-to-Moderate<br>6+ Moderate-to-Severe |
| Module 4 | .98 | 98% | 93% | 3 | 0-2 Little-to-No<br>3-4 Mild-to-Moderate<br>5+ Moderate-to-Severe |

**Table 5.** Percent of participants in each range of concern by diagnostic group

| Concern Ranges | Toddler |  | Module 1 |  | Module 2 |  | Module 3 |  | Module 4 |  |
| --- | --- | --- | --- | --- | --- | --- | --- | --- | --- | --- |
|  | <u>ASD</u> | <u>NonASD</u> | <u>ASD</u> | <u>NonASD</u> | <u>ASD</u> | <u>NonASD</u> | <u>ASD</u> | <u>NonASD</u> | <u>ASD</u> | <u>NonASD</u> |
| Little-to-No | 3.9 | 77.8 | 7.1 | 100 | 1.9 | 56.5 | 9 | 62.5 | 8.5 | 100 |
| Mild-to-Moderate | 6.6 | 22.2 | 17.9 | 0 | 7.5 | 17.4 | 5.7 | 20 | 14.9 | 0 |
| Moderate-to-Severe | 89.5 | 0 | 75 | 0 | 90.5 | 26.1 | 85.2 | 17.5 | 76.6 | 0 |

### Convergent Validity with the ADOS-2

Convergent validity was strong with the ADOS-2 (see Table 6) for the modules that had a significant number of unique participants with an ADOS-2 previously completed during an independent evaluation (i.e., Toddler Module and Module 3). Using Cohen’s (1988) correlation conventions to interpret effect size (i.e., small = .10, medium = .30, large = .50), the strength of the associations was large (Toddler Overall Total and CSS: .74 (p<.001); Module 3 Overall Total: .63 (p<.001); Module 3 CSS: .54 (p<.001)), providing evidence for good convergent validity between the BOSA with the ADOS-2 for those modules.

**Table 6.** Correlations between BOSA Scores and ADOS Scores

|  | <u>BOSA Algorithm Total<sup>+</sup></u> |  |
| --- | --- | --- |
|  | Toddler (n=31) | Module 3 (n=65) |
| Summary Score |  |  |
| ADOS-2 Algorithm Overall Total | .74* | .63* |
| Calibrated Severity Score (CSS) | .74* | .54* |

\*p<.001
<sup>+</sup>Modules 1, 2, and 4 not reported due to limited sample size (i.e., <10 completed ADOS-2s)

### Reliability

Interrater reliability was high (see Table 7), with intraclass correlation coefficients (ICCs) ranging from .90 to .95 across modules. Cross-site reliability between UCLA and CADB coders was .84. For individuals who received multiple BOSA administrations, test-retest reliability was .95. Unlike the ADOS-2 (but similar to most psychometric instruments), we did not look at individual item reliability, as the BOSA is proposed not as a detailed way to collect phenotypic data, but as a brief observation to support the clinical process.

**Table 7.** Reliability: Interrater and Test-Retest

|  | Intraclass Correlation Coefficient |
| --- | --- |
| Toddler Module Interrater | .94** |
| Module 1 Interrater | .93** |
| Module 2 Interrater | .92** |
| Module 3 Interrater | .93* |
| Module 4 Interrater | .90** |
| Cross-Site Interrater | .84** |
| Test-Retest | .95** |
| **p<.01, *p<.05 |  |

## Discussion

The BOSA offers a new option for a remote assessment measure that can be used both in-person and through telehealth. It can be administered by someone without expertise in ASD and scored based on video-recorded observation. Its use of ADOS-2 coding and many of the ADOS-2 materials make it a convenient option for clinicians already trained in the ADOS-2. It could be used as a more accessible and efficient measure to either determine initial risk for ASD, as a level-2 screener to see if a full evaluation with an ADOS-2 or other standardized diagnostic observation is warranted, or in conjunction with other methods (e.g., parent interview and developmental history, behavioral observations) to make a diagnosis. Our intention in creating this measure was to support the clinical and research field to access and deploy the BOSA easily when face-to-face interactions were no longer feasible or safe; therefore, training and materials have been available free-of-charge since conception, with over 20,000 training video views and translation of administration into 5 languages to further facilitate even greater accessibility internationally.

### Improved Feasibility and Accessibility of Brief, Remote Assessment Tools

To improve feasibility of the BOSA for remote administration, materials can be shipped or dropped off to families’ homes or kept locally at community sites. While it would be easier to use whatever toys families have on hand, preliminary testing found that the variation in materials had clear effects on the kinds of play that occurred. Providing materials is also beneficial for families with limited resources. Relatedly, when there are gaps in access to digital technology, devices may be provided for use by the family, or testing can be done at a community site. Clinicians familiar with the ADOS-2 can observe the BOSA live, through telehealth, or on recorded video and complete many of the ADOS-2 codes. This allows for flexibility in each stage of the process: how individuals and families access the materials, where they complete the observation, and when the clinician observes and codes the assessment. Not only has this range of options been ideal for social distancing requirements, but it could also increase accessibility to services for families in more remote or less-resourced areas that may not normally have access to high-quality autism evaluations.

Increasing feasibility and lowering the cost of evaluations can also be improved through the use of briefer, more efficient measures. During the pandemic, the BOSA’s 12-14-minute time limit was likely beneficial in medical centers that were required to follow Center for Disease Control (CDC) guidelines and protocols, as it falls under the threshold for what is considered “close contact”: six feet or less for more than 15 minutes. When social distancing requirements are lifted, assessment options requiring less time from clinicians and families will continue to be desirable to save on clinician time, medical care costs, and burden on individuals and families.

However, using a brief behavioral observation makes collecting parent interview and outside report even more critical. A significant, though relatively small proportion of individuals with ASD fell in the mild-to-moderate concern range, suggesting that the BOSA is likely less precise than the ADOS-2. Additionally, the specificity of the higher modules (i.e., F2 administrations scored with Modules 3 or 4) and the usefulness of the measure in that age group overall may be more limited and less accurate than the ADOS-2, especially when used for first-time diagnoses in adolescents and adults with more subtle symptomatology. The BOSA is unlikely to elicit all of the same skills and behaviors that we would expect to see in an ADOS-2. For example, there is no press for response to joint attention, only some of the socioemotional questions are asked, and a limited set of objects and activities are included, with less likelihood of eliciting restricted and repetitive behaviors. Use of additional measures can clarify whether such symptoms do not exist or were simply not observed during the BOSA.

### Supplemental Testing for a Comprehensive Evaluation

Options for supplemental testing as part of a diagnostic evaluation have been in consideration amongst clinicians during the pandemic and will continue to be a concern as psychological assessment services are offered remotely. Completing cognitive testing remotely can be done but has limitations, particularly for young children and youth with attentional or social difficulties. Solely completing ASD-specific testing does not give a full diagnostic picture and could cause a clinician to misinterpret difficulties or delays related to other factors of development (e.g., when social skills are immature for chronological age, but may be consistent with developmental level). For this reason, understanding an individual’s cognitive functioning is an important factor in putting their social skills and behaviors into context. Some widely used cognitive measures (e.g., Wechsler series) have been made available online, though there are limitations to which tasks can be administered remotely or without using shared materials and manipulatives. Caregivers may be able to act as facilitators in testing cognitive abilities or developmental level for children, though caution should be used in interpretation of results, as parents could unknowingly provide prompting, making it more difficult to determine the child’s true ability level. Interviews, such as the Vineland Adaptive Behavior Scales, Third Edition (Vineland-3; Sparrow et al., 2016), can be conducted over telehealth to aid in obtaining a thorough report on adaptive functioning. With these considerations and limitations in mind, obtaining comprehensive information should be prioritized to get a full and accurate diagnostic picture as assessment services continue to be offered remotely.

### Considerations for Caregiver-Administered Assessment

Using caregivers as the social interactant has unique advantages and disadvantages that are important to consider. Some parents are accustomed to providing a high level of support for their child, especially when it is needed in daily interactions. This is distinctly different from what a trained clinician would do in an ADOS-2: to intentionally hold back to see what the child can do on their own initially, then add in support as needed to end the interaction or activity on a positive, successful note. Because parent support can affect how well a child performs, clinical judgment must be used in determining whether the caregiver’s behaviors may be masking difficulties the individual would have experienced otherwise. Given that it is a brief observation that is not under the clinician’s control, it may not provide as rich of an example to demonstrate to parents how their child fits (or doesn’t fit) autism criteria like the ADOS-2 would. Similarly, because the BOSA is typically administered by caregivers and is so brief, it also does not allow for the observation of ‘emerging’ skills and the degree to which social support from the examiner can buttress the participant’s communication and social behavior. While this can be a limitation of caregiver-led assessment, it can also provide rich information about the parent-child dynamic that is not typically observed in a standard autism evaluation and can be incredibly helpful for individualized recommendations.

### Limitations

Samples were restricted to those that were already being collected before and during COVID-19, as opposed to selecting representative samples with a substantial control group. While the ratio of males to females in this sample is roughly representative of current prevalence data, the results may underrepresent females with possible autism, and the utility of the BOSA for the broader range of females with ASD may not be well-characterized. The broad range of IQs presents an additional limitation, as the Toddler and Module 1 participants have lower mean IQ scores than Modules 2-4, where participants fell within the Average range. Additionally, while we do not see patterns of site differences suggesting ascertainment bias on the results, it is possible that site effects may exist. Not all data was available for all participants, such as ADOS-2 and cognitive scores, or demographic information such as race, ethnicity, and maternal education. Given that so few (i.e., less than 10) participants had a completed ADOS-2 for Modules 1, 2, and 4, correlations between the BOSA and these modules of the ADOS-2 were not statistically significant and could not be interpreted. As with all preliminary studies, but even more importantly with a limited sample, replication studies need to be done. Due to the potentially skewed samples without substantial control groups, recommended cutoffs and ranges of concern should be used with caution and clinical judgment, and reliance on the scores from the BOSA alone should not be used conclusively to make a diagnosis.

### Clinical Utility and Future Applications

While BOSA administration by a caregiver has been more common and feasible during the pandemic with social distancing concerns, use of other test administrators such as a teacher, therapist, or paraprofessional could also greatly improve flexible and timely completion of testing. After COVID-19, therapists or staff can more readily replace caregivers as the administrator, removing the need to disentangle an individual’s independent skills from the support they are receiving. Within early intervention services, therapists or paraprofessionals working with the child could administer this measure to gain information specific to ASD and determine if a full evaluation is warranted. There is continued interest in the use of flexible and remote approaches to ASD assessment, which may be able to improve prompt access to appropriate care for individuals and families. The push to adapt our current approaches to fit social distancing requirements has allowed us to not only continue providing essential services during the pandemic, but also may be seen as a “silver lining” that will allow us to continue expanding the scope and access of care for individuals with ASD long into the future. The pandemic has pushed the field to embrace expansion of more flexible, remote options for ASD assessment services that will continue to shape how we deliver services for years to come.

## Data Availability

All data produced in the present study are available upon reasonable request to the authors.

## Notes

**Conflict of Interest:** Catherine Lord receives royalties from Western Psychological Services (WPS) for the ADOS-2, SCQ and ADI-R. The BOSA is copyrighted by WPS because of its overlap with the ADOS and BOSCC. Deanna Dow, Alison Holbrook, So Hyun Kim, and Catherine Lord are authors of the BOSA, but it is not currently for sale and does not yield any royalties at this time.

### Competing Interest Statement

Catherine Lord receives royalties from Western Psychological Services (WPS) for the ADOS-2, SCQ and ADI-R. The BOSA is copyrighted by WPS because of its overlap with the ADOS and BOSCC. Deanna Dow, Alison Holbrook, So Hyun Kim, and Catherine Lord are authors of the BOSA, but it is not currently for sale and does not yield any royalties at this time.

### Funding Statement

This study was supported by a grant from SFARI (811870, CL).

### Author Declarations

The institutional review board of UCLA gave ethical approval for this work.

## Citations

American Psychiatric Association (APA). (2013). Diagnostic and Statistical Manual of Mental Disorders, 5th Edition (DSM-5). Washington, DC: American Psychiatric Publishing.

Cohen, J. (1988). Statistical Power Analysis for the Behavioral Sciences (2nd ed.). Hillsdale, NJ: Lawrence Erlbaum Associates, Publishers.

Corona, L., Hine, J., Nicholson, A., Stone, C., Swanson, A., Wade, J., Wagner, L., Weitlauf, A., & Warren, Z. (2020). TELE-ASD-PEDS: A telemedicine-based ASD evaluation tool for toddlers and young children. Vanderbilt University Medical Center. https://vkc.vumc.org/vkc/triad/tele-asd-peds

Dahiya, A. V., McDonnell, C., DeLucia, E., & Scarpa, A. (2020). A systematic review of remote telehealth assessments for early signs of autism spectrum disorder: Video and mobile applications. Practice Innovations, 5(2), 150.

Dow, D., Day, T. N., Kutta, T. J., Nottke, C., & Wetherby, A. M. (2020). Screening for autism spectrum disorder in a naturalistic home setting using the systematic observation of red flags (SORF) at 18–24 months. Autism Research, 13(1), 122–133.

Duda, M., Daniels, J., & Wall, D. P. (2016). Clinical evaluation of a novel and mobile autism risk assessment. Journal of Autism and Developmental Disorders, 46(6), 1953–1961.

Gotham, K., Risi, S., Dawson, G., Tager-Flusberg, H., Joseph, R., Carter, A., Hepburn, S., McMahon, W., Rodier, P., Hyman, S., Sigman, M., Rogers, S., Landa, R., Spence, A., Osann, K., Flodman, P., Volkmar, F., Hollander, E., Buxbaum, J., Pickles, A., & Lord, C. (2008). A replication of the Autism Diagnostic Observation Schedule (ADOS) revised algorithms. Journal of the American Academy of Child & Adolescent Psychiatry, 47(6),

Gotham, K., Risi, S., Pickles, A., & Lord, C. (2007). The Autism Diagnostic Observation Schedule: revised algorithms for improved diagnostic validity. Journal of autism and developmental disorders, 37(4), 613–627.

Grzadzinski, R., Carr, T., Colombi, C., McGuire, K., Dufek, S., Pickles, A., & Lord, C. (2016). Measuring changes in social communication behaviors: Preliminary development of the Brief Observation of Social Communication Change (BOSCC). Journal of Autism and Developmental Disorders, 46(7), 2464–2479.

Jang, J., White, S. P., Esler, A. N., Kim, S. H., Klaiman, C., Megerian, J. T., Morse, A., Nadler, C., & Kanne, S. M. (2021). Diagnostic evaluations of autism spectrum disorder during the COVID-19 pandemic. Journal of Autism and Developmental Disorders, 1–12.

Kim, S. H., Thurm, A., Shumway, S., & Lord, C. (2013). Multisite study of new Autism Diagnostic Interview— Revised (ADI-R) algorithms for toddlers and young preschoolers. Journal of Autism and Developmental Disorders, 43(7), 1527–1538.

Lebersfeld, J. B., Swanson, M., Clesi, C. D., & O’Kelley, S. E. (2021). Systematic review and meta-analysis of the clinical utility of the ADOS-2 and the ADI-R in diagnosing autism spectrum disorders in children. Journal of Autism and Developmental Disorders, 1–14.

Lord, C., Holbrook, A., Dow, D., Byrne, K., Grzadzinksi, R., Sterrett, K., Toolan, C., & Kim, S. H. (2020). Brief Observation of Symptoms of Autism (BOSA). plTorrance, CA: Western Psychological Services.

Lord, C., Rutter, M., DiLavore, P.C., Risi, S., Gotham, K, & Bishop, S. (2012a). Autism Diagnostic Observation Schedule, Second Edition (ADOS-2). Torrance, CA: Western Psychological Services.

Lord, C., Luyster, R.J., Gotham, K., & Guthrie, W. (2012b). Autism Diagnostic Observation Schedule, Second Edition (ADOS-2) Manual (Part II): Toddler Module. Torrance, CA: Western Psychological Services.

Luyster, R., Gotham, K., Guthrie, W., Coffing, M., Petrak, R., Pierce, K., Bishop, S., Esler, A., Hus, V., Oti, R., Richler, J., Risi, S., & Lord, C. (2009). The Autism Diagnostic Observation Schedule—Toddler Module: A new module of a standardized diagnostic measure for autism spectrum disorders. Journal of Autism and Developmental Disorders, 39(9), 1305–1320.

Muthén, L. K., & Muthén, B.O. (1998-2011). Mplus User’s Guide (Sixth Edition). Los Angeles, CA: Muthén & Muthén.

Rutter, M., Le Couteur, A., & Lord, C. (2003). Autism Diagnostic Interview—Revised. plTorrance, CA: Western Psychological Services.

Schopler, E., Van Bourgondien, M. E., Wellman, G. J., & Love, S. R. (2010). Childhood Autism Rating Scale– 2nd Edition (CARS-2). Los Angeles, CA: Western Psychological Services.

Schutte, J. L., McCue, M. P., Parmanto, B., McGonigle, J., Handen, B., Lewis, A., Pulantara, I. W., & Saptono, A. (2015). Usability and reliability of a remotely administered adult autism assessment, the Autism Diagnostic Observation Schedule (ADOS) module 4. Telemedicine and e-Health, 21(3), 176–184.

Smith, C. J., Rozga, A., Matthews, N., Oberleitner, R., Nazneen, N., & Abowd, G. (2017). Investigating the accuracy of a novel telehealth diagnostic approach for autism spectrum disorder. Psychological Assessment, 29(3), 245.

Sparrow, S. S., Cicchetti, D. V., & Saulnier, C. A. (2016). Vineland Adaptive Behavior Scales, Third Edition (Vineland-3). San Antonio, TX: Pearson.

Tariq, Q., Daniels, J., Schwartz, J. N., Washington, P., Kalantarian, H., & Wall, D. P. (2018). Mobile detection of autism through machine learning on home video: A development and prospective validation study. PLoS Medicine, 15(11), e1002705.

Wagner, L., Corona, L. L., Weitlauf, A. S., Marsh, K. L., Berman, A. F., Broderick, N. A., Francis, S., Hine, J., Nicholson, A., Stone, C., & Warren, Z. (2021). Use of the TELE-ASD-PEDS for Autism Evaluations in Response to COVID-19: Preliminary Outcomes and Clinician Acceptability. Journal of autism and developmental disorders, 51(9), 3063–3072. https://doi.org/10.1007/s10803

